# Can Machine Learning Algorithms use Contextual Factors to Detect Unwarranted Clinical Variation from Electronic Health Record Encounter Data during the Treatment of Children Diagnosed with Acute Viral Pharyngitis

**DOI:** 10.64898/2026.02.23.26346757

**Authors:** Apollo McOwiti, Yara Neaimeh, Jiaqi Gu, Sana Lalani, Taylor Newsome, Yvonne Nguyen, Meera Subash, Laila Rasmy-Bheket, Susan Fenton

## Abstract

**Rationale, Aims and Objectives:** Unwarranted clinical variation (UCV) in patient care often arises from contextual factors and contributes to increased costs, unnecessary treatments, and deviations from evidence-based practice. Detecting UCV is challenging due to the complexity of care decisions. Current approaches rely on centralized data aggregation and mixed-effects regression, which estimate relative variation but cannot detect absolute variation. Moreover, machine learning (ML) methods leveraging contextual factors for UCV detection are lacking. The objective is to demonstrate the feasibility of ML for identifying absolute UCV using contextual features extracted from electronic health records (EHR) and identify the factors correlated with UCV in treating acute viral pharyngitis in children.

**Methods:** We conducted a retrospective study of pediatric ambulatory visits (ICD-10 J02.8) at an academic health system. The use case focused on unwarranted antibiotic prescriptions for acute viral pharyngitis. We trained ensemble ML models—Random Forest, CatBoost, and Explainable Boosting Machine (EBM)—using encounter-level EHR data. Performance was evaluated using nested cross-validation and AUC metrics. We also compared CatBoost models trained on curated (gold-standard) versus weak labels.

**Results:** All three ML models demonstrated robust performance, with a median AUC of 0.91, using data from 24 clinics, 81 providers, and 122 patients within an academic health system. CatBoost models trained on weak labels exhibited performance comparable to those trained on gold-standard labels. Feature importance analysis indicated that site-level and provider-level case volumes were the most influential predictors, followed by provider credential, years of experience, and encounter type. Notably, lower provider case volumes were associated with a reduced likelihood of inappropriate treatment.

**Conclusions:** Classical ML models can effectively detect absolute UCV using contextual EHR features. Explainable models such as EBM offer interpretability critical for clinical adoption. These findings support ML-based approaches as scalable alternatives to traditional statistical methods for UCV detection without requiring centralized data analysis.

## Introduction

Unwarranted clinical variation(UCV) is common during healthcare provision^1,2^, but has been shown to lead to various problems, including increased cost of care^3,4^, lack of evidence-based care^5^ and poor outcomes^3,6^. There is a great need to identify and to mitigate UCV^7,8^, for example via feedback to providers^8^. One comprehensive definition of UCV is patient care that does not align with patient clinical characteristics, needs or preferences^5^. Unwarranted variation studies conclude that patient clinical factors do not drive the variation measured^2^, meaning that most UCV arises from other contextual factors related to the care, and is a risk posed to the patient by the care system (sites and providers), as opposed to patient behavior. We will refer to these contextual factors as local context factors (LCF). UCV is theorized to arise from the interplay of these LCF^4,5,9^, and in many instances whenever UCV is identified, it is attributed to the provider, even though it is not always obvious what portion of the UCV is due to the provider^10^ or the site.

Identifying UCV is complex and nuanced^5,10^. One analytic framework^5^ introduces three domains of care for analyzing UCV and posits that UCV identification methods involve absolute or relative measurements of processes, costs, resources or outcomes, that can be applied in the three domains of care: capacity, agency or application of evidence base.

Traditional efforts for UCV identification rely on statistical analysis techniques examining patient factors in addition to LCF, such as these examples^11–26^. The analyses use centralized data from participating entities, and the resulting variation is a relative measurement between these entities (regions, sites, hospitals or providers)^4,6,27^. This type of statistical analysis assigns identified special variation (i.e. non-random or common cause variation) to UCV^22,27^, but UCV also arises when there is deviation (for example treatment) from evidence base^5^. Additionally, most of these analyses are based on Wennberg’s small area analysis method^28,29^ that identify region variations^29^, represented as atlases of variation^6,17^.

Whereas such relative UCV measurement are common, there have been few attempts to perform UCV determination using absolute measurements^5^, or deviation. One possible reason for this is the complexity of the care process^5,30^: clinical care process involves multiplicities of choices, processes and decisions a provider makes, and when examining a care pathway, these complexities make it difficult to make an unequivocal judgement on the appropriateness of the care delivered to the patient^30^. One approach to handle such complexities involves the use of decomposition strategy, to decompose a care process and analyze each constituent piece independently^31^. A simple example of this is the case of treating acute viral pharyngitis in children (in this work, we will rely on this particular use case). From the time the patient presents to the provider, several steps are involved, including the interpretation of the treatment guidelines, that can lead to treatment deviation.

It has been shown that adherence to treatment guidelines reduces unwarranted variation, by reducing deviation^1,32^. In the case of acute viral pharyngitis, the Infectious Disease Society of America (IDSA) clinical practice guidelines (CPG) stipulates that no antibiotics should be prescribed to treat acute viral pharyngitis in children^33^. Despite the clear evidence that prescribing antibiotics in such cases is of no benefit to patients and goes against antibiotics stewardship goals^34^, studies show that providers continue prescribe antibiotics as treatment for viral pharyngitis in children^35^. The ISDA guideline on treating viral pharyngitis provides an unequivocal CPG stipulations, and treatment deviation from the ISDA recommendation can be used as an absolute measure of UCV^5^. To test this, instead of using the methods traditionally used to elicit relative UCV, we trained a machine learning (ML) algorithms on electronic health record (EHR) to classify treatment as warranted or unwarranted for the use case of acute viral pharyngitis.

The development of ML algorithms for clinical tasks using EHR data is widespread^22,36–39^ since the study on prediction with EHR data^40^. But for UCV detection, due to clinical care complexities, there are few clinical problems expressed in ways that enables ML discrimination. Additionally, for clinical ML models to trusted by clinicians, they also need to be explainable^41^. Classical ML methods such as logistic regression are explainable out of the box^42^, but deep learning and ensembles models do not provide explanations for the predictions they make^41^. The SHapely Additive exPlanations (SHAP) have been used post-hoc to elicit feature importance from these models^43^. Some ensemble models, such as explainable boosting model (EBM) are designed to be explainable, offering both global and local feature importance scores. The advantage of an EBM model is their ability to provide feature functions for a given feature, isolate any interaction terms between^44^features and examine the contribution of each value group in a categorical feature. Explainable ML for clinical tasks has been developed for various clinical identification tasks, such as rare disease identification^36,45^, patient adherence to guideline^38^, but there is a paucity of reports applying ML to identify UCV using contextual factors derived from EHR data.

Therefore, the study addressed two key questions. First, can machine learning methods, using LCFs derived from routinely collected electronic health record (EHR) data, identify unwarranted clinical variation? Second, which LCFs are associated with inappropriate antibiotic prescribing for acute viral pharyngitis in pediatric patients treated at ambulatory clinics affiliated with an academic medical center?

In this paper, we introduce a ML algorithm for classifying the appropriateness of treatment for acute viral pharyngitis. To overcome the care complexity problem, we apply the decomposition strategy and focus on the treatment phase of viral pharyngitis care. The UCV determination we describe is an absolute measurement of the treatment process. We focus on the treatment offered to the children, to identify treatment deviation from accepted evidence base. We also demonstrate the use standardized features via UCVA Ontology^46,47^ in our dataset. We show that classical ML, especially ensemble methods based on random forest, CatBoost and interpret-ML EBM, result in models with good performance and that are explainable. We also present the five most influential contextual factors associated with unwarranted treatment. To the best of our knowledge, this is the first paper reporting a ML algorithm that identifies UCV using contextual factors from EHR data.

## Methods

### Data source and data elements

Electronic health record (EHR) data were obtained from the BIG-ARC clinical data warehouse (CDW) of UTHealth, which contains structured and unstructured data from UTHealth-affiliated clinics. The CDW data are standardized using the PCORnet Common Data Model^48^. We extracted ambulatory encounter records from January 1, 2021, to December 30, 2024, for children with ICD-10 code J02.8 (acute pharyngitis due to other specified organism). ICD-10 code J02.9 (acute pharyngitis, unspecified) is commonly used for viral pharyngitis; however, J02.8 provides organism-level specificity and typically reflects a confirmed diagnosis in ambulatory care.

For each visit, patient-level variables included sex, race, address, and antibiotic prescriptions. Provider-level variables included national provider identifier (NPI), sex, J02.8 case volume during the study period, total diagnoses in the visit year, encounter type, and encounter year. Site-level variables included J02.8 case volume and total diagnoses for the visit year. Visit date and pseudo-identifiers for patients, providers, and sites were recorded.

### Study population

Children aged 3–19 years with a final ICD-10 diagnosis code of J02.8 at the index encounter were included. The visit date served as the index date. Exclusions were applied for:

1. Antibiotic use at the index date or pre-existing conditions requiring antibiotics (e.g., otitis media, chronic pharyngitis)^49^.
2. Positive group A streptococcal test (rapid or culture) within 3 days after or 7 days before the index date^49^.
3. Absence of clinical notes for the index date or the preceding 7 days.

Chart review of visit notes was approved by the UTHealth Institutional Review Board. Notes from the index date and up to 7 days prior were retrieved for clinical context.

### Local Context Factors Definitions

Contextual factors associated with UCV were defined based on prior research^2^ and standardized using the UCVA ontology^50^. Ontology-based mapping aligned EHR data with UCVA instances.

**Site-level factors:** annual case volumes for all diagnoses and for J02.8, represented numerically and categorically (Low, Medium, High).

**Provider-level factors:** sex(Male, Female); physician indicator (Yes/No); credentials (MD, PA, NP, Other); specialty (Pediatrics, Family Practice, Other); years of experience (numeric, >10 years [Yes/No], and career stage [Early, Mid, Late]); annual case volumes for all diagnoses and J02.8 during the study period.

**Patient-level factors:** sex (male, female, not indicated); race/ethnicity (Caucasian, African American, Hispanic, Other); and socioeconomic status via U.S. Area Deprivation Index (ADI), represented as national percentile (1–100) and state decile (1–10), both encoded numerically and as terciles (Low, Medium, High). National percentile was also classified as federal/state high-needs designation (High/Low Needs)^51,52^.

Although UCV is not directly dependent on patient clinical factors, prior studies suggest associations with ADI and related variables^23,25,53,54^; ADI features were included to assess potential influence on model classification. Encounter year was coded as an ordinal variable (0–3). Table 1 defines all features.

**Table 1.** Site, Provider and Patient Factors, with Feature Labels and Descriptions.

| Factor |  | Description and Type | Mapped UCVA Ontology Instance and Data Property |
| --- | --- | --- | --- |
| Category | Feature Label |  |  |
| Patient | ADI_NATRANK | ADI national rank (numeric) | hasPercentile |
|  | ADI_STATERNK | ADI state decile rank (numeric) | hasDecile |
|  | adi_high_needs | ADI high needs indicator (binary) | ADINeediness_Low ADINeediness_High |
|  | Adi_nat_quartiles | ADI national tercile rank (nominal) | ADINationalTercile_0 ADINationalTercile_1 ADINationalTercile_2 |
|  | Adi_quartiles | ADI state tercile rank (nominal) | ADISTateTercile_0 ADISTateTercile_1 ADISTateTercile_2 |
|  | race_4 | Patient's race (White, Black, Hispanic, Other) |  |
|  | ptsex | Patient sex (F, M, NI) |  |
| Provider | cred | Provider credential (nominal) | ProviderCredential_MD<br>ProviderCredential_NP<br>ProviderCredential_PA<br>ProviderCredential_Other |
|  | enc_type | Encounter type (nominal) | EncounterType_AV EncounterType_ED<br>EncounterType_OA EncounterType_OT<br>EncounterType_TH |
|  | encyear | Encounter year (ordinal) | EncounterYear_0 EncounterYear_1<br>EncounterYear_2 EncounterYear_3 |
|  | exp_bin | experience over 10years (binary) | ProviderExperienceOver10Years_0<br>ProviderExperienceOver10Years_1 |
|  | exp_level | Experience level (nominal) | ProviderExperience_0 ProviderExperience_1<br>ProviderExperience_2 |
|  | exp_years | Years of Experience (numeric) | hasYearsExperience |
|  | phy_case_vol | J02.8 case volume/year (nominal) | ProviderURTICaseVolume_UTP0<br>ProviderURTICaseVolume_UTP1<br>ProviderURTICaseVolume_UTP2 |
|  | phy_case_vol_g | All diagnoses case volume/year (nominal) | ProviderCaseVolume_UTP0<br>ProviderCaseVolume_UTP1<br>ProviderCaseVolume_UTP2 |
|  | phy_case_vol_global | All diagnoses case volume/year (numeric) | hasVolumeNumeric |
|  | phy_ind | MD indicator(binary) | ProviderMDIndicator_0<br>ProviderMDIndicator_1 |
|  | physex | Provider sex(binary) | ProviderGender_UTP0<br>ProviderGender_UTP1 |
|  | Spcl_4 | Provider's specialty (nominal) | ProviderSpecialty_UTP0<br>ProviderSpecialty_UTP1<br>ProviderSpecialty_UTP2 |
| Site | Site_case_vol | J02.8 case volume/year (nominal) | SiteCaseVolume_UTP0<br>SiteCaseVolume_UTP1<br>SiteCaseVolume_UTP2 |
|  | Site_case_vol_g | All diagnoses case volume/year (nominal) | SiteURTICaseVolume_0<br>SiteURTICaseVolume_1<br>SiteURTICaseVolume_2 |
|  | Site_case_vol_global | All diagnoses case volume/year (numeric) | hasVolumeNumeric |

Notes: ADI area deprivation index. The Uri for each instance is: http://sbmi.uth.edu/am/ucva

### Feature Set Definitions

We generated multiple feature sets from the same dataset using different combinations of site, provider, and patient factors to assess their influence on identifying UCV from EHR data. To reduce feature cardinality and enable interoperability, EHR factors were mapped to standardized UCVA ontology instances. This approach addressed two objectives: (1) mitigating high cardinality given the small sample size and (2) supporting cross-institutional comparisons using harmonized UCVA features. Table 1 lists all factors and their corresponding ontology instances.

Features were grouped into nine sets: DD, LCF_DS, ALCF, ALLA, ALLB, ALLC, WEAK_LCF, WEAK_ALLC, and Melogit_DS (Table 2). WEAK_LCF and WEAK_ALLC replicate LCF_DS and ALLC but use weak labels—treatment labels inferred directly from EHR data.

**Table 2.**
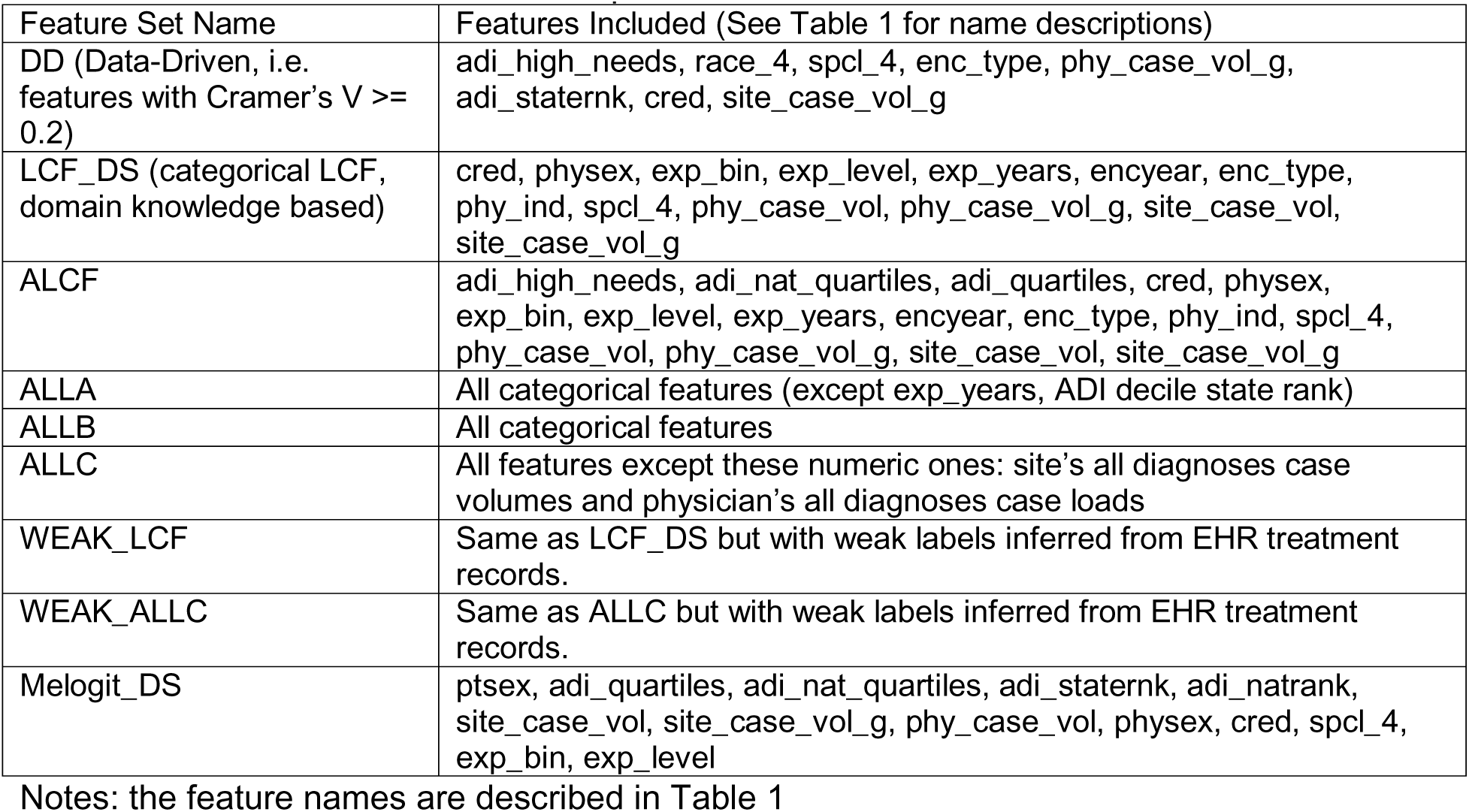
Feature Sets for Model Development.

| Feature Set Name | Features Included (See Table 1 for name descriptions) |
| --- | --- |
| DD (Data-Driven, i.e. features with Cramer's V >= 0.2) | adi_high_needs, race_4, spcl_4, enc_type, phy_case_vol_g, adi_staternk, cred, site_case_vol_g |
| LCF_DS (categorical LCF, domain knowledge based) | cred, physex, exp_bin, exp_level, exp_years, encyear, enc_type, phy_ind, spcl_4, phy_case_vol, phy_case_vol_g, site_case_vol, site_case_vol_g |
| ALCF | adi_high_needs, adi_nat_quartiles, adi_quartiles, cred, physex, exp_bin, exp_level, exp_years, encyear, enc_type, phy_ind, spcl_4, phy_case_vol, phy_case_vol_g, site_case_vol, site_case_vol_g |
| ALLA | All categorical features (except exp_years, ADI decile state rank) |
| ALLB | All categorical features |
| ALLC | All features except these numeric ones: site's all diagnoses case volumes and physician's all diagnoses case loads |
| WEAK_LCF | Same as LCF_DS but with weak labels inferred from EHR treatment records. |
| WEAK_ALLC | Same as ALLC but with weak labels inferred from EHR treatment records. |
| Melogit_DS | ptsex, adi_quartiles, adi_nat_quartiles, adi_staternk, adi_natrank, site_case_vol, site_case_vol_g, phy_case_vol, physex, cred, spcl_4, exp_bin, exp_level |

Notes: the feature names are described in Table 1

These weak-label sets were used to evaluate model performance when trained on unreviewed EHR data.

### Modeling pipeline

Models were developed in Python 3.9 using pandas, scikit-learn, Keras, and interpretML (v0.7.3) for Explainable Boosting Machine (EBM) and CatBoost (v1.2.8). Development was performed in Google Colaboratory. Logistic regression (LR) and random forest (RF) required one-hot encoding for categorical variables.

We conducted three experiments:

1. **Algorithm selection:** LR, RF, EBM, and CatBoost were trained on the ALLC feature set. Nested cross-validation (outer: 10-fold stratified; inner: 10-fold for hyperparameter tuning) was applied to mitigate bias (n = 132). Hyperparameters were optimized via GridSearchCV. Performance was compared using AUC box plots, Friedman Chi-square test, and post hoc pairwise tests. CatBoost and EBM were selected for subsequent experiments. See Table S1 (Supplementary Materials) for model descriptions.
2. **Feature set evaluation:** Six CatBoost models were trained on six feature sets using 10-fold cross-validation with 20 bootstrap repeats. Log loss was used as the loss function and evaluation metric for probability calibration^41,55^.
3. **Weak-label comparison:** Nested cross-validation (10 × 10) compared CatBoost models trained on weak labels (CB_Weak_LCF, CB_Weak_ALLC) versus gold-standard labels (CB_GS_LCF, CB_GS_ALLC).

For interpretability, EBM models (EBM_LCF, EBM_ALCF) provided global feature importance; CatBoost_LCF used SHAP values. The final model was CatBoost trained on LCF_DS. A stratified group split held out 20% (26 samples) for testing; the remaining 106 samples were trained using nested cross-validation (10 × 5). Performance metrics included weighted Precision, Recall, F1-score, AUC (PR and ROC), and Brier score.

### Socioeconomic Data Processing

Patient addresses (street and ZIP) at the visit were used to compute the Area Deprivation Index (ADI). The 2023 Texas ADI dataset was obtained from the Neighborhood Atlas^56^. Addresses were validated via USPS ZIP Code Checker^57^, and GEOIDs were retrieved from U.S. Census data (benchmark = Public_AR_Current; vintage = Current_Current). FIPS codes were constructed by concatenating StateFIPS, CountyFIPS, Tract, and Block; block group was derived from the first Block digit, as ADI is validated at this level.

Using FIPS, we obtained national ADI percentiles and state ADI deciles. Missing FIPS (n = 2) were imputed with the dataset median. For FIPS labeled “GQ-PH” (Group Quarters/Population Housing), ADI was treated as missing (2 cases, 1.5%) and imputed using median state decile and national percentile.

### Accessing Provider Factors by NPI

The EHR encounter data included each provider’s NPI identifier, which we used to retrieve provider information from the NPPES NPI Registry^58^. Using the NPPES API (version 2.1), we queried provider data by passing the NPI identifier as a parameter. From the API response, we extracted the provider’s registration date (used as a proxy for years of experience), credential (e.g., MD, NP, PA, or Other), and specialty (e.g., Pediatrics, Family Practice, or Other).

### Chart Review

Of 38 visits with prescription data, six (15.8%) had positive strep A results and were excluded. Notes for the remaining 32 visits were retrieved from CDW tables and converted to PDF via Python. After applying exclusion criteria, 26 visit notes were retained along with the IDSA CPG and two survey questions (Supplementary Figure S12) to create a REDCap survey.

Two notes were randomly selected for a training survey to familiarize reviewers with note format and relevant CPG definitions. Six physicians reviewed the remaining notes, with each note assessed by at least two reviewers; disagreements were adjudicated by a third reviewer. Reviewers had one month to complete surveys. Data were exported to Excel and analyzed in Stata.

### Multi-level Clustering Assessment

Provider-within-site clustering was evaluated using Stata’s mixed-effects logistic regression (melogit). A random-effects model was fitted to calculate intraclass correlation coefficients (ICC) for site and site/provider clusters. Fixed effects were introduced starting with feature set DD, but models for DD, LCF, and ALLC did not converge. To address this, alternative feature combinations were tested, and Melogit_DS achieved convergence with ICC ≈ 0, indicating negligible clustering.

Consequently, logistic regression with robust standard errors and generalized estimating equations (GEE) at the site level was applied. Sensitivity analyses across all three models assessed robustness.

### Statistical Analysis

Analyses were performed using Stata vBE19.5 and Python’s *statsmodels*. Normality was assessed via QQ-plots and Shapiro–Wilk tests. Group comparisons used Pearson’s Chi-square for categorical variables (Fisher’s exact when cell counts <5) and Mann–Whitney U for continuous variables. Associations were quantified with Cramer’s V.

Due to unmet homogeneity assumptions, nonparametric tests were applied: Friedman Chi-square and Wilcoxon Signed Rank for post hoc comparisons. Clustering analyses used Stata’s *melogit* (mixed-effects), *logit* (robust errors at site/provider), and *xtgee* (GEE). Sensitivity analyses of odds ratios were performed with *esttab*.

Inter-rater reliability was assessed using Cohen’s Kappa, and ICD-10 J02.8 coding accuracy was evaluated with *cii*. Statistical significance was set at α = 0.05.

## Results

### Encounter Selection

A total of 132 encounters coded as ICD-10 J02.8 were identified across 24 clinics, 80 providers, and 122 patients. Of these, 112 visits (no-treatment group) involved no antibiotic prescription, representing 19 sites, 71 providers, and 104 patients. The treatment group included 38 visits with antibiotics; six (15.8%) had positive strep A results and were excluded from chart review.

Chart review was performed on 32 treatment-group visits; exclusions included three with pre-existing conditions (9.4%) (ENT surgery, throat infection), two lacking sufficient documentation (6.3%), and one with chronic pharyngitis (3.1%), leaving 26 visits for appropriateness assessment. After review, 20 visits were classified as unwarranted antibiotic prescriptions. These 20 visits involved 10 sites, 14 providers, and 18 patients.

For machine learning tasks, 26 visits (20%) were held out for evaluation; all 132 encounters were included in multilevel regression analyses. Site/provider and patient feature summaries are provided in Tables 3 and 4.

**Table 3.**
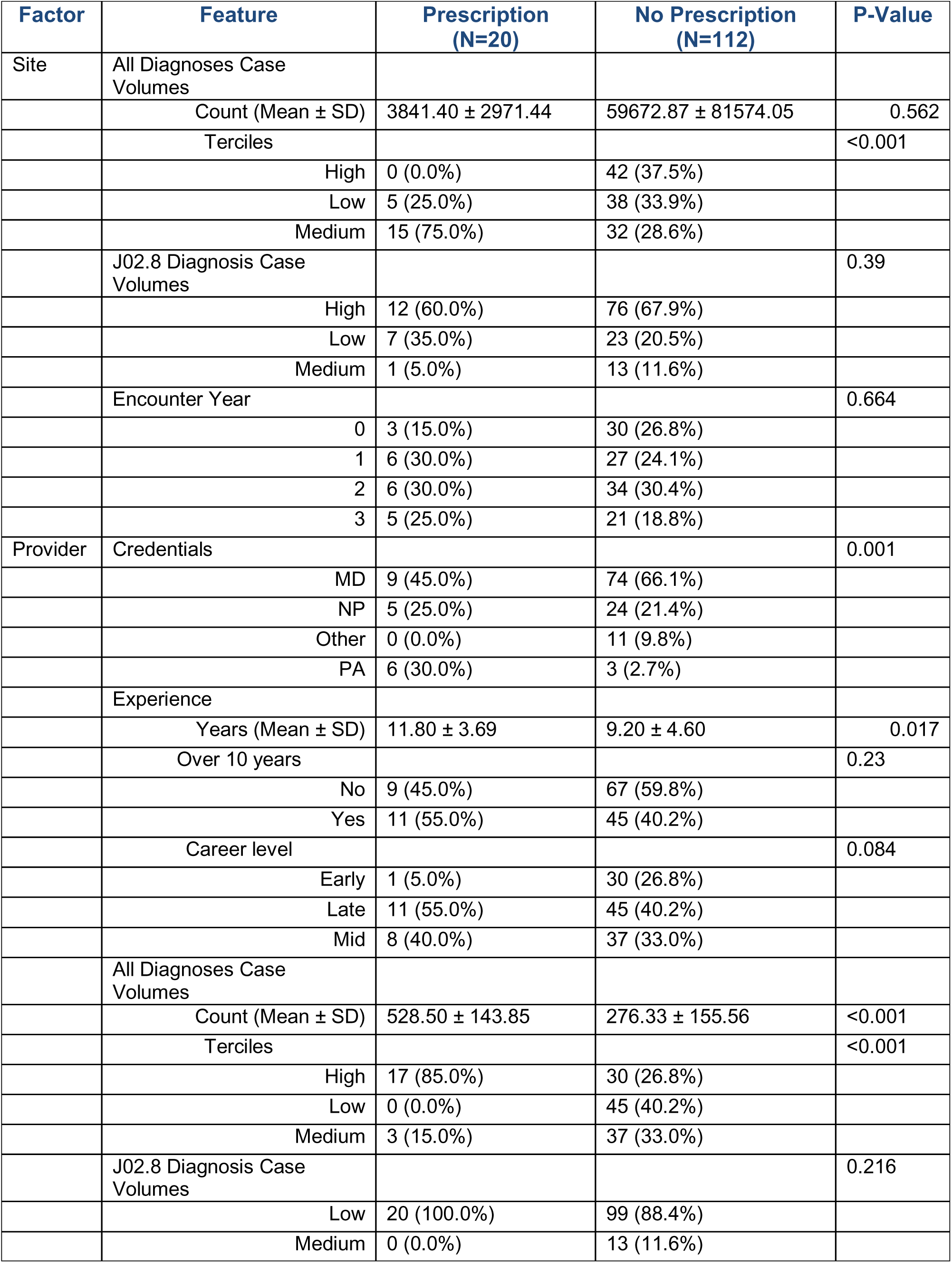
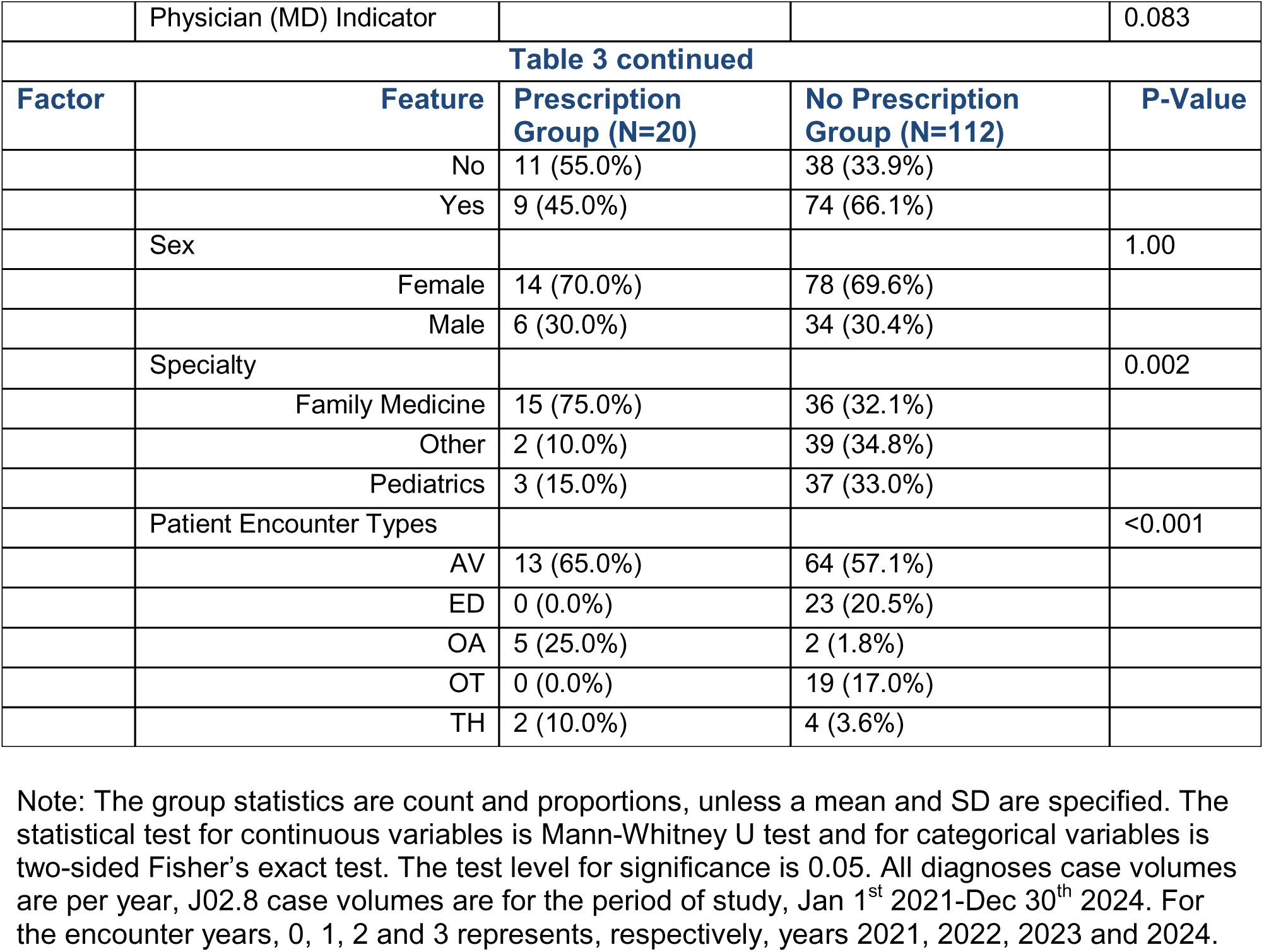
Site and provider factors grouped by prescription and no prescription samples.

| Factor | Feature | Prescription<br>(N=20) | No Prescription<br>(N=112) | P-Value |
| --- | --- | --- | --- | --- |
| Site | All Diagnoses Case Volumes |  |  |  |
| | Count (Mean ± SD) | 3841.40 ± 2971.44 | 59672.87 ± 81574.05 | 0.562 |
|  | Terciles |  |  | <0.001 |
|  | High | 0 (0.0%) | 42 (37.5%) |  |
|  | Low | 5 (25.0%) | 38 (33.9%) |  |
|  | Medium | 15 (75.0%) | 32 (28.6%) |  |
|  | J02.8 Diagnosis Case Volumes |  |  | 0.39 |
|  | High | 12 (60.0%) | 76 (67.9%) |  |
|  | Low | 7 (35.0%) | 23 (20.5%) |  |
|  | Medium | 1 (5.0%) | 13 (11.6%) |  |
|  | Encounter Year |  |  | 0.664 |
|  | 0 | 3 (15.0%) | 30 (26.8%) |  |
|  | 1 | 6 (30.0%) | 27 (24.1%) |  |
|  | 2 | 6 (30.0%) | 34 (30.4%) |  |
|  | 3 | 5 (25.0%) | 21 (18.8%) |  |
| Provider | Credentials |  |  | 0.001 |
|  | MD | 9 (45.0%) | 74 (66.1%) |  |
|  | NP | 5 (25.0%) | 24 (21.4%) |  |
|  | Other | 0 (0.0%) | 11 (9.8%) |  |
|  | PA | 6 (30.0%) | 3 (2.7%) |  |
|  | Experience |  |  |  |
| | Years (Mean ± SD) | 11.80 ± 3.69 | 9.20 ± 4.60 | 0.017 |
|  | Over 10 years |  |  | 0.23 |
|  | No | 9 (45.0%) | 67 (59.8%) |  |
|  | Yes | 11 (55.0%) | 45 (40.2%) |  |
|  | Career level |  |  | 0.084 |
|  | Early | 1 (5.0%) | 30 (26.8%) |  |
|  | Late | 11 (55.0%) | 45 (40.2%) |  |
|  | Mid | 8 (40.0%) | 37 (33.0%) |  |
|  | All Diagnoses Case Volumes |  |  |  |
| | Count (Mean ± SD) | 528.50 ± 143.85 | 276.33 ± 155.56 | <0.001 |
|  | Terciles |  |  | <0.001 |
|  | High | 17 (85.0%) | 30 (26.8%) |  |
|  | Low | 0 (0.0%) | 45 (40.2%) |  |
|  | Medium | 3 (15.0%) | 37 (33.0%) |  |
|  | J02.8 Diagnosis Case Volumes |  |  | 0.216 |
|  | Low | 20 (100.0%) | 99 (88.4%) |  |
|  | Medium | 0 (0.0%) | 13 (11.6%) |  |
|  | Physician (MD) Indicator |  |  | 0.083 |
| **Table 3 continued** |  |  |  |  |
| **Factor** | **Feature** | **Prescription Group (N=20)** | **No Prescription Group (N=112)** | **P-Value** |
|  | No | 11 (55.0%) | 38 (33.9%) |  |
|  | Yes | 9 (45.0%) | 74 (66.1%) |  |
|  | Sex |  |  | 1.00 |
|  | Female | 14 (70.0%) | 78 (69.6%) |  |
|  | Male | 6 (30.0%) | 34 (30.4%) |  |
|  | Specialty |  |  | 0.002 |
|  | Family Medicine | 15 (75.0%) | 36 (32.1%) |  |
|  | Other | 2 (10.0%) | 39 (34.8%) |  |
|  | Pediatrics | 3 (15.0%) | 37 (33.0%) |  |
|  | Patient Encounter Types |  |  | <0.001 |
|  | AV | 13 (65.0%) | 64 (57.1%) |  |
|  | ED | 0 (0.0%) | 23 (20.5%) |  |
|  | OA | 5 (25.0%) | 2 (1.8%) |  |
|  | OT | 0 (0.0%) | 19 (17.0%) |  |
|  | TH | 2 (10.0%) | 4 (3.6%) |  |

Note: The group statistics are count and proportions, unless a mean and SD are specified. The statistical test for continuous variables is Mann-Whitney U test and for categorical variables is two-sided Fisher's exact test. The test level for significance is 0.05. All diagnoses case volumes are per year, J02.8 case volumes are for the period of study, Jan 1<sup>st</sup> 2021-Dec 30<sup>th</sup> 2024. For the encounter years, 0, 1, 2 and 3 represents, respectively, years 2021, 2022, 2023 and 2024.

**Table 4.**
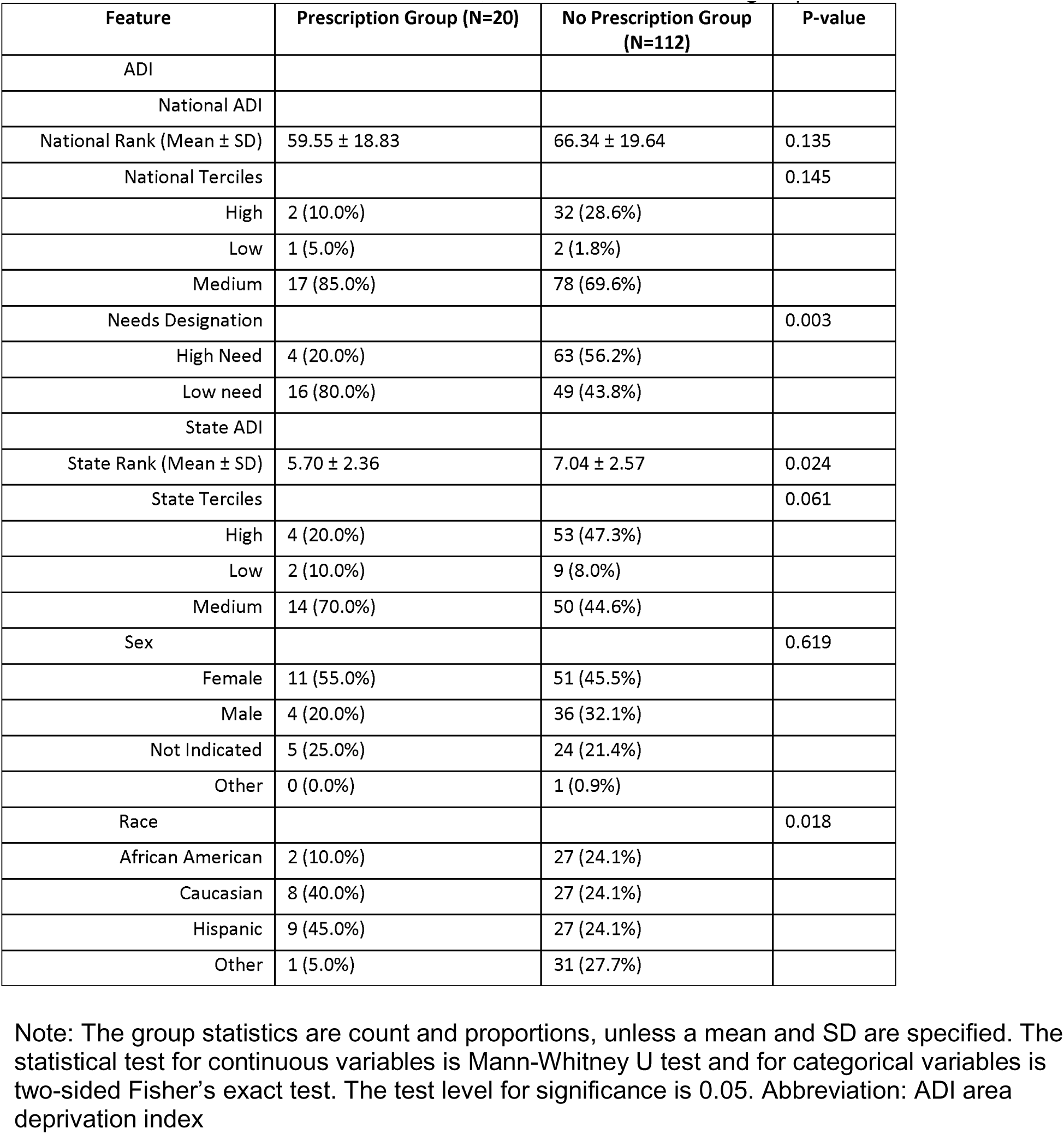
Patient non-clinical factors of ADI, sex and race between groups.

### Classification of appropriate treatment from EHR data

#### Model Selection

Four machine learning models—logistic regression (LR), random forest (RF), Explainable Boosting Machine (EBM), and CatBoost—were trained on the ALLC feature set to classify inappropriate treatment. RF achieved the highest stability and performance (median AUC 0.90, IQR 0.14), followed by EBM (0.89, IQR 0.13) and CatBoost (0.89, IQR 0.15). LR performed lower (mean AUC 0.85, SD 0.15), Figure 1. Ensemble models outperformed LR due to their ability to capture nonlinear feature interactions.

**Figure 1:**
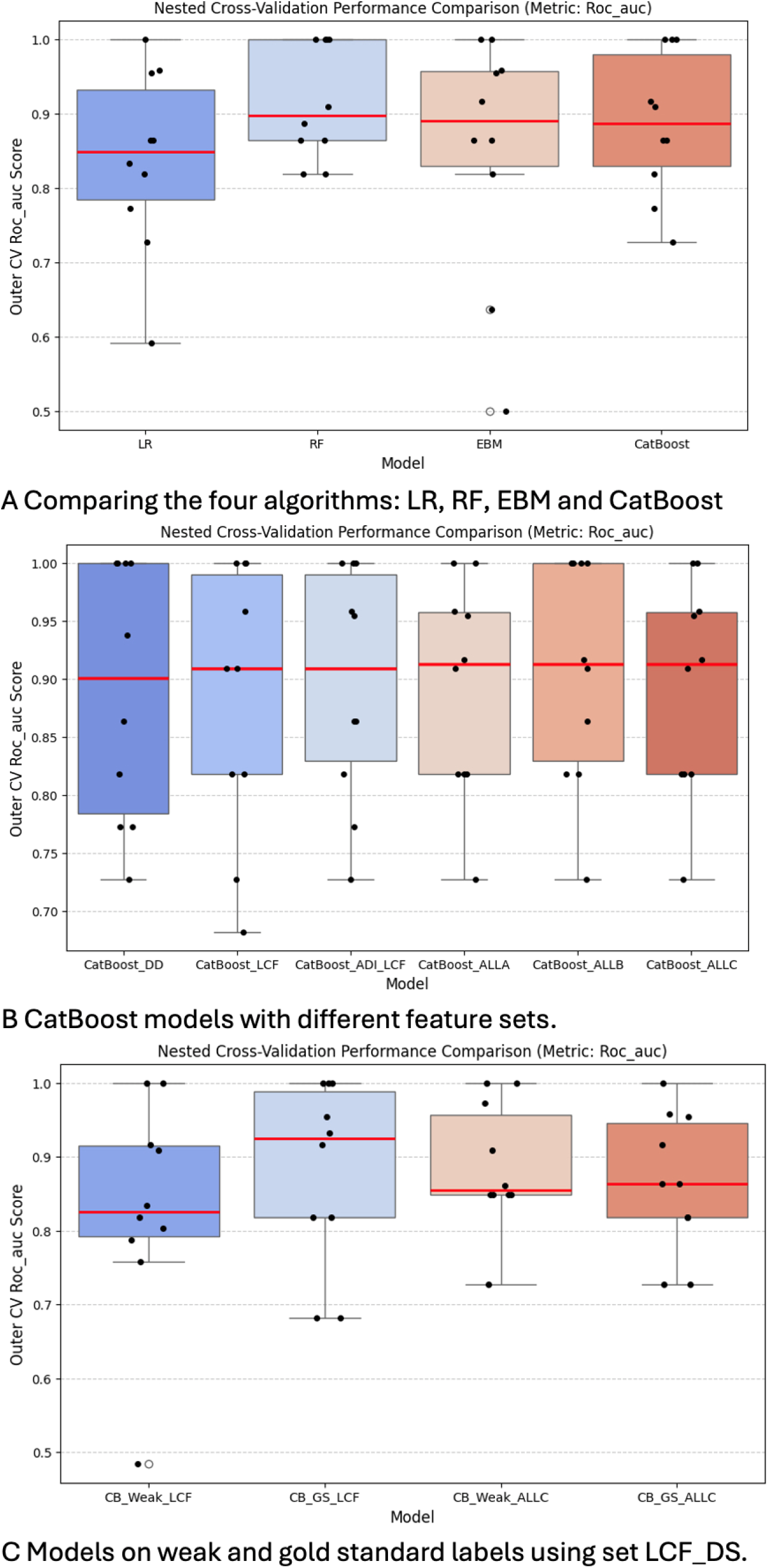
Comparing ML classifiers. Part A, models based on four different algorithm (Logistic Regression (LR), Random Forest (RF), CatBoost and Explainable Boosting Machines (EBM)) trained on the ALLC feature set to classify inappropriate treatment. RF achieved the highest stability and performance (median AUC 0.90, IQR 0.14), followed by EBM (0.89, IQR 0.13) and CatBoost (0.89, IQR 0.15). LR performed lower (mean AUC 0.85, SD 0.15). Only LR and RF show statistically significant difference in AUC performance. Part B CatBoost models trained on different feature sets of same samples. CatBoost_DD has the lowest median AUC (0.90 IQR 0.22), the rest have same AUC (0.91), with varying IQR values. CatBoost_ALLA is the most stable model. Friedman test (Chi2 3.62, p-value 0.61) shows there is no significant difference between the mean AUC performance of these models. Part C, CatBoost comparing use of weak labels using two different feature sets. On feature set LCF, Model CB_GS_LCF shows better AUC performance (AUC 0.0.92, IQR 0.0.17) than CB_Weak_LCF (AUC 0.838 IQR 0.12). On feature set ALLC, model CB_GS_ALLC (AUC 0.86 IQR 0.13) is slightly better than model CB_Weak_ALLC (AUC 0.85 IQR 0.13).

Friedman Chi-square test indicated significant differences among models (χ² = 8.667, p = 0.034). Post hoc Wilcoxon tests with Bonferroni correction showed only LR vs RF differed significantly (p = 0.0078, α = 0.0083). Given comparable performance and complementary strengths, EBM and CatBoost were selected for further experiments.

#### Feature Set Selection

CatBoost, optimized for categorical data, was trained on six feature sets (DD, LCF_DS, ALCF, ALLA, ALLB, ALLC). Median AUCs were: CatBoost_DD (0.90, IQR 0.22), CatBoost_LCF (0.91, IQR 0.17), CatBoost_ALCF (0.91, IQR 0.16), CatBoost_ALLA (0.91, IQR 0.14), CatBoost_ALLB (0.91, IQR 0.17), and CatBoost_ALLC (0.91, IQR 0.14). CatBoost_DD performed worst; ALLA was most stable among the top five, figure 1(bottom diagram).

All models used identical samples and cross-validation folds, isolating feature set impact. Friedman test (χ² = 3.62, p = 0.61) showed no significant performance differences.

CatBoost_LCF was selected for further experiments because it relies solely on local context factors (site and provider volumes, credentials, sex, experience, encounter type/year, physician indicator, specialty), all mapped to UCVA ontology.

#### Weak Labels vs Gold Standard Labels

We compared CatBoost models trained on weak labels (CB_Weak_LCF, CB_Weak_ALLC) and gold-standard labels (CB_GS_LCF, CB_GS_ALLC), Figure 1. For LCF features, CB_GS_LCF outperformed CB_Weak_LCF (AUC 0.92, IQR 0.17 vs. 0.84, IQR 0.12). For ALLC features, CB_GS_ALLC (AUC 0.86, IQR 0.13) slightly exceeded CB_Weak_ALLC (AUC 0.85, IQR 0.13).

Wilcoxon tests showed no significant differences between weak and gold-standard models (LCF: W = 13.0, p = 0.30; ALLC: W = 13.0, p = 0.55). Friedman test across all four models confirmed no significant AUC differences (χ² = 2.83, p = 0.42). These findings suggest weak labels inferred from EHR data perform comparably to chart-reviewed labels.

#### Feature Importance

Figure 2 presents global feature importance for models EBM_LCF (A), EBM_ALCF (B), and CatBoost_LCF (C). For EBM models, the y-axis lists features, and the x-axis shows log-odds contributions. In EBM_LCF, the top predictors were provider and site case volumes (phy_case_vol_g, site_case_vol_g), provider credential (cred), experience level (exp_level), and sex (physex). For EBM_ALCF, the leading features were provider and site case volumes, national ADI quartiles (adi_nat_quartiles), high-needs ADI (adi_high_needs), and provider specialty (spcl4). CatBoost_LCF used SHAP values; the most influential features were provider case volume, site case volume, site J02.8 case volume, provider experience, and encounter type (ENC_TYPE). Case volume features dominated across models, with provider case volume being most influential. Most other features clustered near zero, indicating few strong predictors.

**Figure 2:**
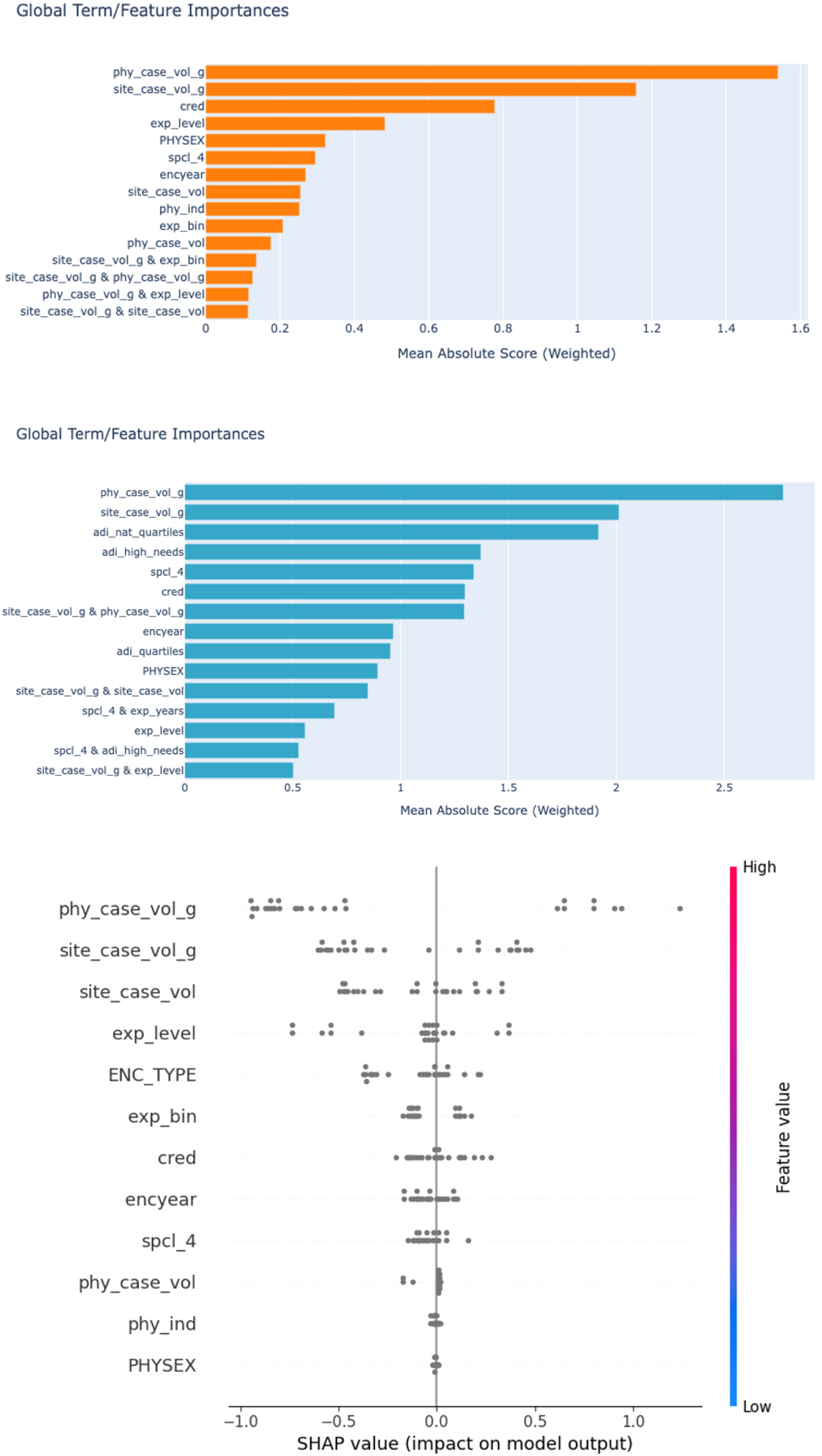
Global feature importance summaries for models EBM_LCF (A) and EBM_ALCF(B) and SHAP values for model CatBoost_LCF(C). For EBM models, the y-axis is the feature, the x-axis is the weighted mean absolute score, which is the logs odds of a feature’s contribution to the model prediction. For model CatBoost_LCF, feature importance is shown using SHAP values. In SHAP plot color scheme, gray shows the contribution of a feature that is not encoded in SHAP’s low value(blue) or high value(red) color scheme. Each dot represents an observation and those to the left (negative SHAP value) of the vertical line are negatively associated and those to right are positively associated with the positive outcome (antibiotics treatment), respectively.

Partial dependence plots for select features from models EBM_LCF and EBM_ALCF (Figure 3) illustrate log-odds contributions: three of the top predictors and patient ADI, to investigate how the model depends on the patient ADI. Scores near zero indicate no effect (and reference categories are near zero as well); positive scores increase, and negative scores decrease the likelihood of antibiotic prescribing. For provider case volume (3A), low and high volumes reduced and increased scores, respectively, compared to medium volume. For site case volume (3B), high volume decreased and medium slightly increased scores relative to low volume. For provider credential (3C), NP decreased scores, while PA and Other slightly increased scores compared to MD. The difference was statistically significant (p-value < 0.0001), but pairwise Chi-squared analyses with Bonferroni correction of provider credential indicated that only two comparisons reached statistical significance: PA versus MD (adjusted Bonferroni p = 0.0008) and PA versus Other (adjusted Bonferroni p = 0.0362) (Table S3 in Supplementary Materials).For patient ADI (3D), high-needs ADI reduced scores relative to low-needs ADI.

**Figure 3.**
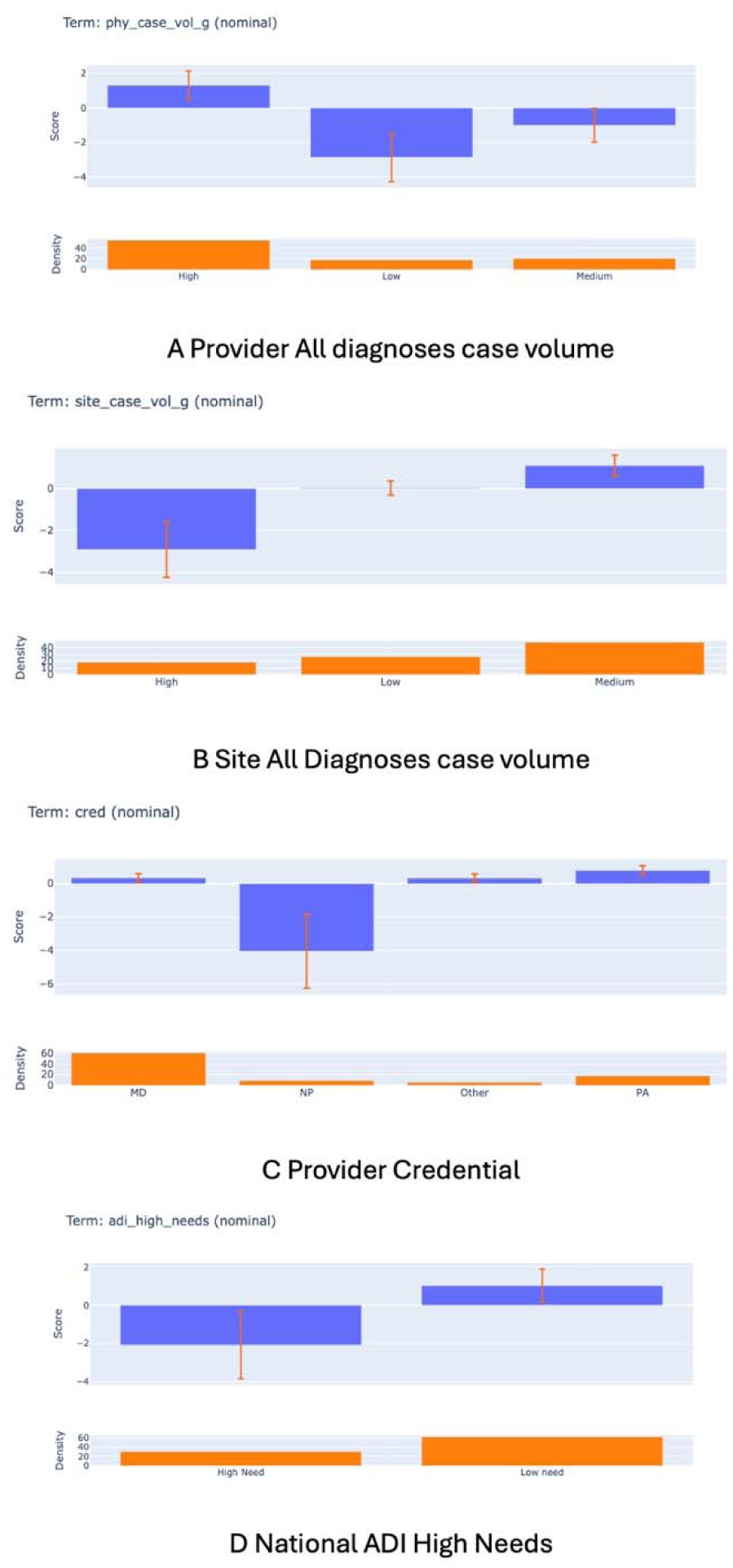
Feature function plots for select features from models EBM_LCF (A, B, C), EBM_ALCF(D). In each diagram, the top figure shows the logs odds score of a feature on the y-axis and the feature value on the x-axis. The vertical red bar is an error bar. The bottom diagram shows the density or distribution function of a feature, and for categorical variables shows the proportion of each category group within the feature. The positive outcome is antibiotics prescription. A: Provider all diagnosis case volume plot. With Medium as reference, Low case volume decreases the likelihood of the positive outcome. B: Site all diagnosis case volume plot. With Low case volume as reference, High site volume decreases the likelihood of outcome. C: Provider Credential plot. With MD as reference, NP decreases the likelihood of positive outcome. D: Patient ADI neediness plot. With Low Need as reference, High Need decreases the likelihood of outcome. See Supplementary Figure S3, and Figure S4 for the other top 5 function plots for model EBM_LCF and EBM_ALCF, respectively.

#### Feature Importance Over Time

We evaluated whether the importance of key features varied over time. Figure 4 displays function plots for the two most influential predictors—provider and site all-diagnosis case volumes—across encounter years 0–1 (2021–2022) and 2–3 (2023–2024). For provider case volume (phy_case_vol_g), shown in panels A (years 0–1) and B (years 2–3), patterns remained consistent: with medium case volume as the reference, low case volume was associated with a reduced score for the positive outcome. For site case volume (site_case_vol_g), panel C (years 0–1) and panel D (years 2–3) similarly demonstrated that High case volume decreased the score for the positive outcome compared with Low case volume.

**Figure 4:**
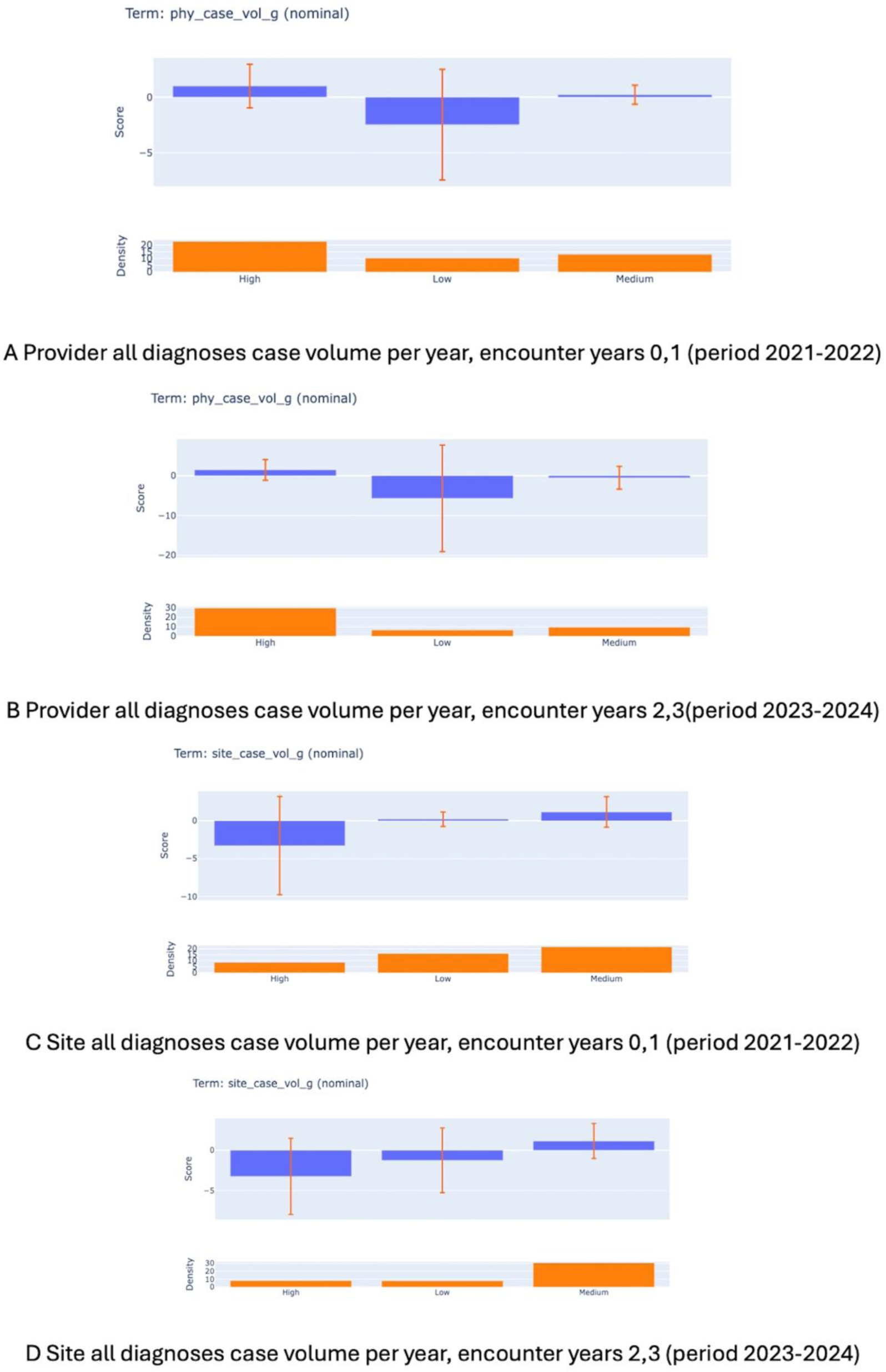
Top two important features, provider all diagnoses and site all diagnoses case volumes during the periods 2021-2022 and 2023-2024. The wide error bars (red vertical line) are because of small sample size causing more uncertainty. For provider case volume (phy_case_vol_g), A (years 0–1) and B (years 2–3), with Medium case volume as the reference, Low case volume was associated with a reduced score for the positive outcome. For site case volume (site_case_vol_g), C (years 0–1) and D (years 2–3) similarly demonstrated that High case volume decreased the score for the positive outcome compared with Low case volume.

#### Evaluation Performance

The final model was a CatBoost classifier trained on the LCF_DS feature set and evaluated on a held-out test set of 26 samples. The model achieved an AUC of 0.89 and a PR-AUC of 0.79. Weighted average performance metrics were Precision (0.85), Recall (0.79), and F1-score (0.81). The Brier score was 0.14, indicating good probability calibration.

### Feature Coefficients with Mixed Effects Analysis

#### Multi-level Cluster Analysis

Ensemble machine learning algorithms leverage nonlinear feature relationships to improve predictive performance compared with logistic regression. To assess clustering effects, we implemented two-level hierarchical models: providers nested within sites and patients nested within providers. When only random effects were modeled, substantial clustering was observed, as indicated by high intraclass correlation coefficients (ICC): site-level ICC = 0.52 (95% CI, 0.16–0.86) and site-provider ICC = 0.93 (95% CI, 0.64–0.99) (Supplementary Figure S6). Model convergence was achieved only when the Melogit_DS feature set was included as fixed effects, which reduced ICC values to near zero (Supplementary Figure S7). Specifically, site-level ICC was 1.02 × 10⁻³³ (SE 2.18 × 10⁻¹□) and site-provider ICC was 1.26 × 10⁻³³ (SE 2.23 × 10⁻¹□), indicating no residual clustering after covariate adjustment. Likelihood ratio testing provided no evidence of improved fit for the mixed-effects logistic model compared with standard logistic regression (Wald χ² = 22.69; p = 0.117).

Given these findings, we fitted standard logistic regression with site-clustered robust standard errors and generalized estimating equations (GEE). Odds ratios were nearly identical across the three approaches (mixed-effects, robust SE logistic regression, and GEE).

#### Logistic Regression with Site-clustered Robust SE and GEE

For this model, Wald χ² = 1887 (p < 0.00001), and pseudo-R² = 0.54. Model performance included specificity of 96%, sensitivity of 85%, and overall classification accuracy of 93%. After adjustment for all covariates, provider and site case volumes were not significantly associated with the outcome (Supplementary Figures S8, S8b). Significant predictors included provider male sex (OR 7.00; 95% CI, 1.20–40.85; p = 0.03), specialty categorized as “Other” (OR 0.05; 95% CI, 0–1.02; p = 0.05), >10 years of experience (OR 383.54; 95% CI, 8.40–17,515.95; p < 0.001), and mid-level experience (OR 101.07; 95% CI, 3.82–2673.35; p = 0.01). The GEE model with small-sample correction yielded similar log-odds estimates (Supplementary Figure S9).

Sensitivity analyses comparing the three logistic approaches (mixed-effects with two-level clustering, site-clustered robust SE, and GEE) demonstrated consistent odds ratios and identical significant predictors across models (Supplementary Figure S10), indicating that results were robust to model specification.

### Unwarranted Treatment Gold Standard Determination

Chart review was conducted to establish gold-standard labels and validate the accuracy of the J02.8 diagnosis code. All encounters coded as J02.8 were reviewed for confirmation. Agreement between coded and confirmed diagnoses was 84%, yielding a positive predictive value (PPV) of 84% (95% CI, 69–94%). Of 38 visits reviewed, six (15.8%) were miscoded; these cases had positive Group A streptococcal results and should have been coded as J02.0.

Inter-rater reliability (IRR) for classification of warranted versus unwarranted antibiotic treatment demonstrated moderate agreement, with Cohen’s kappa = 0.76 and percentage agreement of 88.5%. Among 26 charts reviewed for unwarranted treatment, six (23.1%) were reclassified as warranted, leaving 20 cases confirmed as unwarranted. This corresponds to an estimated unwarranted treatment rate of 15.15% (n = 132) for acute viral pharyngitis across the academic clinic network.

## Discussion

Care for patients with pharyngitis involves multiple phases, each presenting opportunities for unwarranted clinical variation (UCV). Identifying UCV is inherently complex, requiring nuanced decisions across these steps^5^. When researchers decompose this complexity into simpler components, those components become amenable to classical machine learning (ML) methods. Deep learning might be able to learn patterns despite that complexity^40^, with large datasets, but most real-world clinical datasets are small^44^, making classical approaches more practical.

To demonstrate ML applicability for UCV detection, we selected acute viral pharyngitis—a use case with clear, binary treatment expectations—and focused on the treatment phase of care. This decomposition allowed us to frame the problem as a yes/no classification task suitable for ML. Using EHR-derived features, we showed that ensemble models—Random Forest, CatBoost, and Explainable Boosting Machine (EBM)—can effectively classify treatment variation. Importantly, EBM models provide inherent explainability, enabling clinicians to interpret and address unwarranted variations.

The AUC performance of Random Forest, CatBoost, and EBM models was comparable; however, algorithm selection should consider factors beyond accuracy. EBM models were less stable than CatBoost or Random Forest but offer interpretability. CatBoost did not outperform Random Forest, yet it is optimized for categorical data and requires minimal preprocessing. Statistical tests indicated no significant performance differences among these models, except between logistic regression and Random Forest. Therefore, model choice should weigh practical advantages such as stability, interpretability, and preprocessing requirements.

To evaluate the final model (CatBoost trained on LCF_DS, though similar results could be obtained with Random Forest or EBM), we used a held-out dataset to ensure generalizability. Given the small sample size and class imbalance, achieving a stratified split with identical distribution was challenging. Because our goal was to detect deviations—the minority class—we applied class weights to favor this class. This approach increased recall at the expense of precision, which aligns with UCV identification priorities. We consider the cost of false positives (e.g., incorrectly flagging treatment as UCV) less critical than missing true UCV cases.

When testing different feature sets, the prediction task and folds remained constant. A model trained on the LCF_DS feature set (local context factors) achieved the best performance, and adding additional features did not improve results. This finding aligns with evidence that UCV is primarily driven by contextual factors^2,5^. Due to the limited sample size, we could not include all 28 previously identified LCFs^2^. With a larger dataset and sites in diverse locations, future work could incorporate these factors to assess their influence.

We identified five factors with the greatest feature importance: case volumes (site-level for all diagnoses and J02.8 diagnosis, and provider-level for all diagnoses), provider credential, experience level, and encounter type. Case volumes were the top three influential factors. Providers with lower case volumes were less likely to prescribe inappropriate treatment compared to those with higher volumes. While this may seem intuitive—fewer patients allowing more time per case—it contrasts with findings in other settings where higher case volumes are associated with better outcomes^59,60^. Those studies, however, looked at surgery volumes, where higher volume of surgeries was associated with more provider experience and facility capacity. For ambulatory setting, further research is needed to understand the underlying reasons for this association.

An interesting finding was that providers with NP credentials were less likely to prescribe inappropriate treatment compared to MDs (The NP vs MD difference did not meet statistical significance), and early-career providers were less likely than late-career providers to do so. This may reflect differences in decision-making: experienced providers might rely on clinical judgment to “cover bases” preemptively^49^, whereas less experienced clinicians may adhere more closely to clinical practice guidelines. In our chart review, when antibiotics were prescribed despite negative strep A results, providers documented these decisions as preemptive.

An additional finding was that patients from high-needs ADI areas were less likely to receive inappropriate treatment compared to those from low-needs ADI areas. This aligns with systematic reviews reporting higher rates of antibiotic overprescription among high SES (i.e. low needs ADI) groups^61^. Prior studies suggest that patient pressure can lead to inappropriate antibiotics prescribing^62^, which may indicate that low-needs patients exert more influence than high-needs patients. Alternatively, in this over-use scenario, clinicians may be more mindful of resource limitations when treating high-needs ADI populations^4,28^. Further research is needed to clarify these dynamics.

The finding that models trained with weak labels performed comparably to those trained with gold-standard labels is noteworthy. Creating gold-standard labels is resource-intensive and time-consuming; therefore, achieving similar performance without manual annotation offers significant advantages. However, this benefit may be limited to simple use cases such as ours, and further research is needed to evaluate its applicability in more complex scenarios.

The mixed-effects model indicated that intraclass correlation coefficients (ICC) for site-level and provider-level clustering were near zero, suggesting minimal clustering. Covariates in the model appeared to account for site and provider contributions, leaving no unexplained within-cluster correlation. Due to the small cluster sizes (24 sites; median of 2 patients per provider), random intercept variances were poorly estimated^63^. Logistic regression with robust standard errors adequately modeled covariate relationships, and sensitivity analysis showed similar log-odds and p-values across models. However, because this analysis relied on patient features not directly relevant to UCV, we do not recommend interpreting the regression coefficients.

This study has three main limitations. First, the dataset was small. While this did not affect classical machine learning methods, it limited the use of deep learning approaches, which typically require large datasets. Deep learning models might have captured factors influencing UCV that classical algorithms could not identify.

Second, we could not perform external evaluation because datasets from other sites were unavailable. This limitation is common in UCV research, as sites are unlikely to share data that reveal operational dynamics. Even with external data, generalizability would remain challenging because UCV is driven by site-specific contextual factors. A model developed for one local context may not perform well elsewhere. We think UCV identification efforts are always going to be localized: the presence of UCV during care can be uncomfortable^8,64^. However, the absence of external validation should not be viewed as a disadvantage. Models that standardize features using the UCVA ontology enable legitimate comparisons of UCV rates across disparate sites, even without direct generalization.

Third, our dataset may contain bias. Because the study is retrospective, some encounters—such as telephone calls—may not have been documented. For example, during chart review, we identified one note indicating that a provider called a patient to report a negative throat culture result after initiating pre-emptive treatment. If the result was negative, the treatment would have been warranted and excluded from the dataset. Additionally, providers may omit documentation of treatment decisions influenced by patient preference, which would also remove those encounters from analysis.

## Conclusion

This study demonstrates that machine learning methods can effectively identify instances of unwarranted clinical variation (UCV) using routinely captured electronic health record (EHR) encounter data. Ensemble-based algorithms outperformed traditional logistic regression for this classification task, achieving median AUC values between 0.89 and 0.91. Importantly, models trained on weak labels inferred from EHR data performed comparably to those trained on gold-standard annotated labels.

Site-level and provider-level case volumes emerged as the most influential factors associated with inappropriate antibiotic prescribing for pediatric acute viral pharyngitis, and these associations persisted across all four study years. Additionally, nurse practitioners were less likely than physicians to provide inappropriate treatment, and lower provider experience was associated with reduced unwarranted prescribing.

## Data Availability

No data was produced for this study

## Acknowledgement

I appreciate the work of the following people. Susan G. Hilsenbeck, PhD, Professor, Baylor College of Medicine, for advice on various issues. Sebastian Shrager, MD, Department of Pediatrics, McGovern Medical School, UTHealth Center at Houston for assisting with the chart review.

